# Fronto-limbic and Thalamocortical Network Alterations after COVID-19 Recovery: a Multimodal MRI Study

**DOI:** 10.64898/2026.05.19.26353613

**Authors:** Sapna S Mishra, Rohit Misra, Gwenaëlle Douaud, Bharat Biswal, Tapan K. Gandhi

## Abstract

**Background:** Persistent neurological and cognitive symptoms following SARS-CoV-2 infection point to long-term alterations in brain structure and function. The thalamus, orbitofrontal cortex, and limbic networks are particularly susceptible to inflammatory and neurovascular stressors. However, the relationship between cortical, white-matter, and thalamocortical alterations in post-COVID syndrome remains unclear.

**Methods:** 76 COVID-19 recovered participants (CRPs) and 51 healthy controls (HCs) underwent multimodal MRI comprising T1-weighted structural, diffusion, and resting-state functional acquisitions. Grey-matter morphology was assessed using voxel-based morphometry (VBM), white-matter microstructure using tract-based spatial statistics (TBSS), and thalamocortical functional connectivity (TC-FC) using seed-based analyses from major thalamic nuclei. Results were evaluated both across the groups (HC vs. CRP) and after stratifying CRPs by hospitalisation status (HC vs. Non-hospitalized patients (NHPs) vs. Hospitalized patients (HPs)).

**Results:** No group-level grey-matter differences were observed between HCs and CRPs; however, HPs showed localized volume loss in the orbitofrontal and frontal-pole cortices (*p_FWE_ <* 0.05). TBSS revealed widespread microstructural abnormalities, including reduced fractional anisotropy and mean diffusivity across association and commissural tracts (*p_corr_ <* 0.05), with regional increases in mode of anisotropy indicating selective loss of crossing fibres (*p_corr_ <* 0.05). Resting-state analyses revealed increased TC-FC from the mediodorsal thalamic nucleus to anterior cingulate, parietal, and occipital cortices (*p_corr_ <* 0.05), while differences in pulvinar and ventrolateral nuclei were not significant (*p_corr_ >* 0.05).

**Conclusions:** Our findings indicate that COVID-19 recovery is associated with enduring alterations in fronto-limbic and thalamo-cortical circuits, most prominently in individuals with severe infection. Convergent structural and functional changes involving the orbitofrontal cortex and mediodorsal thalamus suggest network-specific reorganisation that may underpin persistent cognitive and affective symptoms of post-COVID syndrome.

## 1. Introduction

A substantial proportion of individuals recovering from SARS-CoV-2 infection continue to experience persistent neurological and cognitive symptoms. These long-lasting effects, collectively termed post-COVID syndrome (PCS) or long COVID, include fatigue, attention and memory problems, sleep disturbance and mood alterations [1, 2, 3]. Such symptoms suggest that COVID-19 can leave lasting imprints on the brain, but the mechanisms remain unclear.

Neuroimaging studies indicate that the brain is not spared from the systemic and inflammatory sequelae of infection. Longitudinal analyses from the UK Biobank have shown cortical thinning and grey-matter loss in orbitofrontal and limbic regions even after mild disease, implicating fronto-limbic circuits in long-term effects [4, 5]. Diffusion MRI studies have revealed widespread white-matter alterations suggestive of neuroinflammatory or demyelinating processes [6, 7, 8, 9, 10]. Resting-state fMRI investigations have reported abnormal connectivity in networks supporting attention, salience and sensorimotor integration [11, 12, 13]. Yet, these findings remain heterogeneous, often limited to a single imaging modality and without systematic stratification by disease severity.

Our previous work on the same Indian cohort has consistently identified subcortical and limbic involvement. Diffusion tractography revealed microstructural changes in the fornix and cingulum [14]. Fixel-based analysis showed fibre-specific alterations in limbic tracts [15]. A multimodal study demonstrated convergent abnormalities in the basal ganglia and limbic system [16], while behavioural assessments confirmed persistent fatigue, anxiety and attention deficits [17]. Together, these results point to enduring disruption of fronto-limbic and thalamo-cortical systems that mediate motivation, cognition and sensory regulation.

Building on this background, we conducted a multimodal MRI analysis combining three complementary techniques in COVID-19 recovered participants (CRPs) and healthy controls (HCs). First, we used voxel-based morphometry (VBM) on T1-weighted images to test whether cortical or subcortical grey-matter volumes differ between groups, and whether these effects vary with severity. Structural changes in orbitofrontal or limbic regions may index inflammatory injury or neurovascular compromise underlying cognitive and emotional symptoms. Second, we employed tract-based spatial statistics (TBSS) on diffusion MRI to examine white-matter microstructure across major tracts. This approach tests whether pathways connecting the thalamus, frontal and parietal cortices (critical for cognitive control and sensory integration) show persistent alterations following recovery.

Finally, we investigated thalamocortical functional connectivity (TC-FC) using resting-state fMRI. The thalamus acts as a central hub integrating sensory, cognitive and affective information [18] and is particularly vulnerable to hypoxia, inflammation and vascular injury [19, 20, 21]. Recent work has identified altered thalamocortical coupling in post-COVID patients, especially those with fatigue and cognitive slowing [22, 23]. Leitner and colleagues reported increased thalamic connectivity with frontal and parietal cortices, suggesting thalamic dysregulation as a neural substrate of PCS-related fatigue. Examining TC-FC therefore provides a hypothesis-driven means to test whether functional reorganisation parallels the structural and microstructural disruptions observed in the same cohort.

This study addressed three questions: (1) do recovered COVID-19 patients show lasting alterations in grey- or white-matter architecture?, (2) are these abnormalities dependent on the severity of the acute infection?, and (3) do thalamocortical networks exhibit functional reorganisation that mirrors or compensates for these structural changes? We hypothesised that CRPs, particularly those hospitalised during infection, would exhibit reduced orbitofrontal volume, altered white-matter integrity in association and thalamo-cortical tracts, and abnormal thalamo-cortical connectivity. By integrating these modalities, we sought to provide a unified picture of how COVID-19 affects the brain’s structural and functional architecture and to identify potential imaging markers of PCS.

## 2. Methods

### 2.1. Subject recruitment

For this cross-sectional study, we recruited two groups of subjects: (i) HCs and (ii) CRPs. We used a database of 2358 COVID-19 patients who were treated at a local hospital to recruit the CRPs. 40% of these patients required oxygen therapy, 22% received continuous positive airway pressure (CPAP), and 14% needed intubation. Two weeks after they tested PCR negative, we contacted 100 patients from this database for this study. Additionally, we also contacted 126 CRPs and 100 HCs who were recruited from open calls for participation in the study.

Participants in the CRP group were included based on the following criteria: (i) age greater than 18 years, (ii) laboratory confirmation of COVID-19 infection through a positive RT-PCR test, and (iii) documented recovery from the infection, evidenced by a subsequent negative RT-PCR report obtained within six months prior to MRI acquisition. The inclusion criteria for healthy controls (HCs) were: (i) age greater than 18 years and (ii) no history of COVID-19 infection at the time of scanning. Symptomatic individuals were required to provide a negative RT-PCR report to confirm eligibility as HCs. Exclusion criteria for both groups included: (i) a history of neurological or psychiatric illness, (ii) prior brain injury or neurosurgical intervention, and (iii) for the CRP group, having required life-support measures (e.g., mechanical ventilation) as part of their COVID-19 treatment.

MRI data were acquired from 51 HCs (mean age = 31.98 ± 8.97 years; 12 females) and 76 CRP participants (mean age = 32.38 ± 11.52 years; 19 females) (see Figure S1). Prior to analysis, all images underwent modality-specific quality assessment. Based on these evaluations, 2 participants were excluded from the T1-weighted MRI analysis, 4 from diffusion MRI analysis, and 19 from functional MRI analysis (details provided in subsequent sections). Additionally, 67 CRPs (mean age = 31.12 ± 10.78 years; 16 females) provided information regarding their clinical history during the infection, and 59 (mean age = 30.85 ± 10.52 years; 15 females) reported symptoms experienced following recovery, as confirmed by a negative RT-PCR result.

All data were collected under the purview of the Indian Institute of Technology Delhi, and all MRI acquisitions were performed at Mahajan Imaging Centre, New Delhi, in compliance with the regulations of the Institutional Review Board (IRB). The study protocol was approved by the Institute Ethics Committee of the Indian Institute of Technology Delhi. Written informed consent was obtained from all participants prior to the collection of imaging and behavioral data. The dataset will be made available upon reasonable request to the corresponding authors, subject to institutional data-sharing policies.

### 2.2. Stratification based on infection severity

Of the 76 COVID-19 recovered participants (CRPs) enrolled, 67 consented to provide detailed clinical histories from the time of infection. Participants were subsequently stratified into Hospitalized Patients (HPs) or Non-Hospitalized Patients (NHPs) based on whether they required inpatient care during their COVID-19 illness, with hospitalization used as a proxy for severe disease. HPs received interventions such as continuous positive airway pressure (CPAP), supplemental oxygen, or continuous clinical monitoring, whereas NHPs reported comparatively mild symptoms (e.g., fever, cough, anosmia, ageusia, fatigue) that did not necessitate hospital admission. In total, 21 participants (39.48 ± 11.95 years; 6 females) were categorized as HPs, and 46 (27.30 ± 7.72 years; 10 females) as NHPs. These subgroups were then used to investigate the impact of severity-based stratification on MRI-derived measures.

### 2.3. Magnetic resonance imaging

MRI data were acquired using a 3 T GE Discovery MR750w scanner equipped with a 32-channel head coil. T1-weighted structural images were obtained using a 3D fast BRAVO sequence with the following parameters: flip angle = 12°, inversion time (TI) = 450 ms, field of view (FOV) = 256 × 256 mm², slice thickness = 1.0 mm, and 152 sagittal slices, resulting in 1 mm isotropic resolution.

Diffusion-weighted images were acquired using a spin-echo echo-planar imaging (SE-EPI) sequence with Repetition Time (TR) = 16000 ms, Echo Time (TE) = 79.6 ms, and a matrix size of 256 × 256 × 78. The voxel size was 1 × 1 × 2 mm³ with a slice thickness of 2 mm. Diffusion encoding was performed along 30 directions with a b-value of 1000 s/mm² using anterior–posterior phase encoding. Additionally, four non-diffusion-weighted (b = 0 s/mm²) T2-weighted reference volumes were acquired.

Resting-state functional MRI (rs-fMRI) scans were obtained using a gradient echo-planar imaging (GE-EPI) sequence with the following parameters: TR = 2000 ms, TE = 30 ms, flip angle = 90°, FOV = 240 × 240 mm², matrix size = 64 × 64, and slice thickness = 3 mm (38 slices; voxel size = 3.75 × 3.75 × 3 mm³). Each rs-fMRI acquisition lasted for 800 s (13 min 20 s). During the resting-state scan, participants were instructed to remain still, keep their eyes open, and avoid falling asleep.

### 2.4. Voxel-based morphometry

The T1-weighted MRI were used to study long-term differences in tissue morphology at the voxel-level in the CRPs compared with the HCs. After checking the quality of the images by visual inspection, MRIs of 75 CRPs (32.55 ± 11.79 years, 18F) and 50 HCs (31.68 ± 8.29 years, 12F) were included in the group comparison (HC vs CRP). Next, to study the impact of severity-based stratification on abnormalities in tissue morphology, we included the T1-w MRIs of 20 HPs (38.85 ± 11.90 years, 5 Female) and 46 NHPs (27.17 ± 7.73 years, 11 Female) along with the 50 HCs.

#### 2.4.1. Data processing

Voxel-based morphometry (VBM) is a computational neuroimaging technique used to detect voxel-level differences in brain tissue volumes [24]. In this study, T1-weighted anatomical scans were preprocessed using the SPM12 toolbox (http://www.fil.ion.ucl.ac.uk/spm/) in MATLAB (MathWorks Inc., Natick, MA, USA).

Tissue segmentation was then performed to obtain gray matter (GM), white matter (WM), and cerebrospinal fluid (CSF) maps. A study-specific template was constructed using DARTEL, a fast diffeomorphic image registration algorithm, providing improved spatial normalization by iteratively aligning individual tissue maps to an averaged group template [25].

Each subject’s segmented maps were non-linearly warped to the DARTEL template and subsequently normalized to the standard Montreal Neurological Institute (MNI) space using an affine transformation. These normalized images were resampled to 1.5 mm isotropic resolution. To account for local volume changes introduced by the nonlinear deformation, Jacobian modulation was applied. Finally, contraction-modulated images were smoothed with an 8 mm full-width at half-maximum (FWHM) Gaussian kernel.

#### 2.4.2. Statistical analysis

The smoothed contraction-modulated maps obtained from the VBM analysis were compared using a two-sample t-test, where age, sex, and total intracranial volume (TIV) were used as covariates of no interest. Regions with significant differences in grey matter volume were identified by using a height threshold of *p_unc_ <* 0.001, and corrected for multiple comparisons at a family-wise error (FWE) of *p_FWE_ <* 0.05.

To investigate the effect of severity-based stratification on the VBM maps, we conducted an Analysis of Covariance (ANCOVA), modelling severity as a three-level factor. We added age, sex, and TIV as covariates of no interest. Post-hoc comparisons were conducted using two-sample t-tests. For both ANCOVA and post-hoc tests, we used a voxel-level threshold of *p_unc_ <*0.001 and corrected for multiple comparisons at FWE of *p_FWE_ <* 0.05.

### 2.5. Tract-based spatial statistics

The DWIs were used to study the long-term differences in white matter microstructure in CRPs compared with HCs. Among the 76 CRPs and 51 HCs whose DWIs were acquired, we had to reject the images from 3 CRPs and 1 HC due to poor quality (detected by visual inspection). Overall, we included 50 HCs (31.68 ± 8.79 years, 12F) and 73 CRPs (32.04 ± 11.48 years, 18F) in this analysis. Further, after quality check, 20 HPs (38.85 ± 11.9 years, 5F) and 44 NHPs (27.36 ± 7.86 years, 9F) were included in the study to investigate the effect of infection severity on white matter microstructure.

#### 2.5.1. Data processing

Tract-Based Spatial Statistics (TBSS) is a specialized voxel-wise method for the analysis of diffusion MRI data, designed to enhance sensitivity and anatomical precision in assessing white matter microstructure [26]. By aligning individual diffusion maps to a common white matter skeleton, TBSS circumvents inter-subject anatomical variability, enabling robust comparisons of diffusion metrics such as fractional anisotropy (FA), mean diffusivity (MD), and mode of anisotropy (MO) [27, 28]. The MO metric characterizes the nature of anisotropy in a voxel with values ranging from -1 (planar anisotropy; e.g. regions with crossing/kissing fibres) to +1 (linear isotropy; e.g. regions with one principal fibre orientation) [28].

The dMRI preprocessing pipeline included denoising using Marchenko Pastur Principal Component Analysis (MP-PCA) to reduce thermal noise using MRtrix3 [29]. Gibbs ringing artifacts were also corrected employing MRtrix3. Susceptibility-induced distortions near air-tissue interfaces were addressed using FSL’s TOPUP [30], leveraging synthetic reversed-phase encoding images generated using the SynB0-DISCO algorithm [31]. Eddy current and motion-related distortions were corrected with FSL’s EDDY tool [32]. Brain extraction was performed using FSL’s BET [33].

For the TBSS analysis, individual FA maps were derived from preprocessed DWIs using the diffusion tensor model (FSL’s DTIFIT tool) [26]. These FA maps were then non-linearly registered to MNI space using the FMRIB58_FA template. A mean FA map was created from all aligned FA maps and subsequently thresholded at FA *>* 0.2 to define the white matter skeleton, which reflects the central trajectory of major fibre bundles common across subjects. Each subject’s FA data were then projected onto the mean skeleton using a local maximum search perpendicular to the skeleton tract [26]. FA maps were eroded to reduce residual artifacts, generating a white matter skeleton, and aligned to the MNI space. Similarly, the MD and MO maps were also registered to the MNI space and projected onto the white matter skeleton. Subject-wise skeletonized FA, MD, and MO maps were used for statistical comparisons.

#### 2.5.2. Statistical analysis

To compare the FA, MD, and MO values across the HC and CRP cohorts, we used permutation testing, utilizing the “randomise” tool by FSL [34]. Testing was conducted using 5000 random permutations of the classification. The p-values were thresholded at *p_corr_ <*0.05 using the Threshold-Free Cluster Enhancement (TFCE) method [35].

Similarly, the effect of COVID-19 severity-based stratification was assessed using the “randomise” tool. A design matrix with regressors for each subgroup was supplied to generate contrasts. The p-values were thresholded at *p_corr_*< 0.05 using the TFCE method.

### 2.6. Thalamocortical functional connectivity

Using the resting-state fMRI, we studied the long-term effect of COVID19 on thalamocortical functional connectivity in CRPs as compared to HCs. From the total set of participants, we excluded 15 CRPs and 4 HCs from the fMRI study due to high motion artifacts (maximum framewise translation or rotation *>* 2*mm*), leading to a sample size of 61 CRPs (32.02 ± 11.01 years, 14F) and 47 HCs (32.30 ± 9.23 years, 11F). The effect of severity based stratification on thalamocortical functional connectivity was studied using 47 HCs (32.20 ± 9.23 years, 11F), 14 HPs (41.21 ± 19.59 years, 5F), and 39 NHPs (27.54 ± 8.22 years, 7F).

#### 2.6.1. Data processing

Resting-state fMRI data were preprocessed using the SPM toolbox. Initially, we performed slice-time correction followed by rigid-body realignment to correct for head motion by estimating six transformation parameters (three translations and three rotations). The mean functional images of each subject were co-registered to the respective T1-weighted anatomical scan using a mutual information-based method [36]. Tissue probability maps obtained from segmentation of the anatomical images were used to normalize the anatomical images to MNI standard space non-linearly. The resulting deformation fields were applied to the functional data to facilitate voxel-wise group comparisons [37]. Normalized functional volumes were smoothed using an 8 mm full-width at half-maximum Gaussian kernel [38]. Mean signals from white matter and cerebrospinal fluid, the six motion parameters, and the global mean signal were regressed out as nuisance signals [39]. Finally, temporal band-pass filtering between 0.01 and 0.1 Hz was applied to the functional time series [40, 41].

Thalamocortical functional connectivity (TC-FC) supports the integration of sensory, motor, and cognitive information between the thalamus and cerebral cortex. As a central hub, the thalamus modulates key processes such as sensation, attention, and perception, making TC-FC critical for brain function. Alterations in TC-FC have been linked to various neurological and psychiatric conditions, frequently correlating with deficits in cognition and sensory processing.

Using the preprocessed resting-state fMRI data, we assessed TC-FC originating from the three largest nuclei of the thalamus, namely the Medio-Dorsal Nucleus (MDN), Pulvinar Nucleus (PVN), and the Ventro-Lateral Nucleus (VLN) [42, 43]. For each nucleus, two spherical seed regions (radius = 8mm) with centres corresponding to the MNI coordinates of the nuclei in the right and left thalamus were analysed [43]. The MNI coordinates (in mm) for the MDN, PVN, and the VLN were [±6, 16, 9], [±15, 31, 5], and [±15, 13, 9], respectively [43, 42]. The representative time series were extracted by averaging the BOLD signal within each seed and correlated voxel-wise with the rest of the brain. Pearson correlation coefficients were used to determine the functional connectivity between the thalamic nuclei and the regions of the brain. The correlation coefficients were Fisher *z*-transformed to enable parametric statistical inference.

#### 2.6.2. Statistical analysis

The TCFC maps obtained from the seed-to-voxel analysis were compared voxel-wise across HCs and CRPs using a two-sample t-test, where age and sex were used as covariates of no interest. Regions with significant differences in TCFC were identified by using a height threshold of *p_unc_ <* 0.001, and corrected for multiple comparisons at a family-wise error (FWE) of *p_FWE_ <* 0.05.

To investigate the effect of severity-based stratification on the TCFC, we conducted an Analysis of Covariance (ANCOVA), modelling severity as a three-level factor. We added age and sex as covariates of no interest. Post-hoc comparisons were conducted using two-sample t-tests. For both ANCOVA and post-hoc tests, we used a voxel-level threshold of *p_unc_ <* 0.001 and corrected for multiple comparisons at a family-wise error (FWE) of *p_FWE_ <* 0.05.

## 3. Results

### 3.1. Cohort demographics and characteristics

A total of 127 participants were included and successfully scanned. Demographic characteristics of the cohorts considered in the analyses are summarized in Table 1. Among the CRPs, 67 provided detailed clinical histories for the acute illness phase, on the basis of which 46 were classified as NHPs and 21 as HPs. The HP group comprised CRPs who had required hospitalization and who had received remdesivir (10/21), oxygen supplementation (10/21), or corticosteroids (3/21). The most frequently reported acute COVID-19 symptoms among CRPs were fever (85%) and cough (70%), followed by body ache (64%), loss of smell (45%), chills (37%), loss of taste (37%), breathing difficulties (34%), nausea (19%), and weakness (15%).

**Table 1:** Demographic details of the participants recruited for this study. The age distribution is represented using the “mean ± standard deviation” values.

|  | COVID-19 Recovered Participants |  |  | Healthy Controls |  |  |
| --- | --- | --- | --- | --- | --- | --- |
|  | Number | Age (years) | Sex | Number | Age (years) | Sex |
| Total Recruited | 76 | $32.38 \pm 11.52$ | 19 F/ 57 M | 51 | $31.98 \pm 8.96$ | 12 F/ 39 M |
| Shared COVID-19 infection details | 67 | $31.12 \pm 10.78$ | 16 F/ 51 M | NA | | |
| Shared experience with post-COVID symptoms | 59 | $30.85 \pm 10.52$ | 15 F/ 44 M | 27 | $30.33 \pm 7.85$ | 6 F/ 21 M |

Post-acute symptom data were available for 59 CRPs (30.85 ± 10.52 years; 15 females) who consented to report their post-COVID experience and any persistent complaints. In this subgroup, fatigue (68%) was the most commonly reported post-COVID symptom, followed by unrefreshing sleep (46%) and reduced attention (44%). Additional frequently reported symptoms included achy muscles (44%), headache (41%), and achy joints (39%).

### 3.2. No significant group differences detected using voxel-based morphometry

VBM was employed to quantify grey matter volume for each participant at the voxel level. Group comparisons between CRPs and HCs using two-sample t-tests did not reveal any significant clusters surviving FWE correction at *p_FWE_ <* 0.05.

### 3.3. Severity stratification reveals localised morphological differences

The effect of severity-based stratification was tested using a three-level ANCOVA. Pair-wise post-hoc tests were conducted to discern which groups were significantly impacted. We found a significant effect of severity-based stratification on voxel-wise tissue volume (*p_FWE_ <* 0.05) (see supplementary Figure S2). Post-hoc pairwise comparisons among HCs, NHPs, and HPs demonstrated significant reductions in gray matter volume in HPs relative to both HCs (*p_FWE_ <* 0.05, cluster size = 1043 voxels, peak MNI coordinates: [30, 49.5, -16.5], peak *t*-score = 4.68) and NHPs (*p_FWE_ <* 0.05, cluster size = 835 voxels, peak MNI coordinates: [13.5, 49.5, -25.5], peak *t*-score = 5.03). These differences were localized primarily in the left orbitofrontal cortex and frontal pole (Figure 1). No significant differences were observed between HCs and NHPs. VBM analysis indicates that while no gross grey matter volume differences were observed between CRPs and HCs, severity-based stratification reveals regional cortical volume reductions, specifically in the orbitofrontal cortex and frontal pole in hospitalized patients.

**Figure 1:**
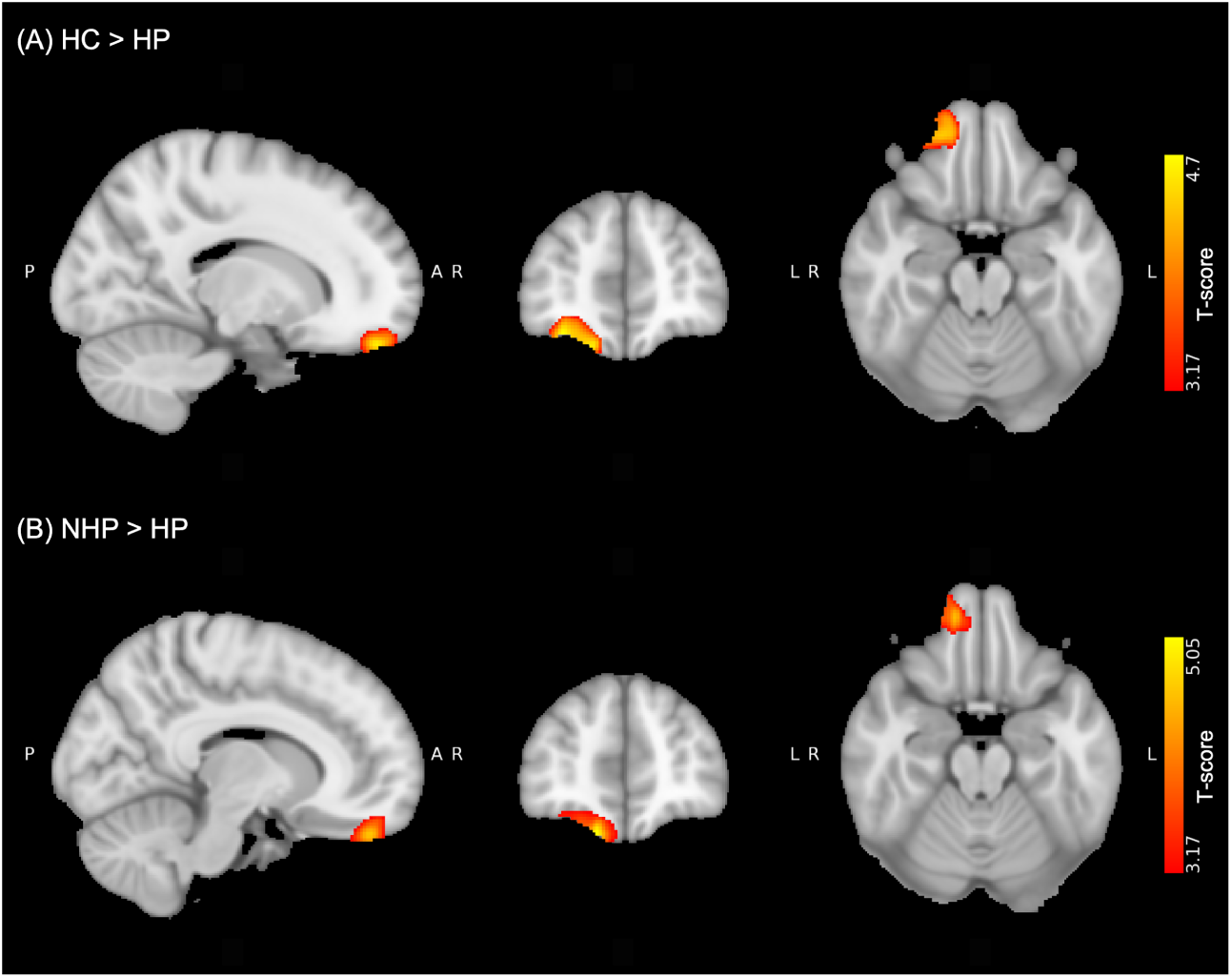
Significant clusters that highlight the effect of severity-based stratification on voxel-based grey matter volume. (A) Post-hoc comparison of Healthy Controls (HCs) and Hospitalized Patients (HPs) highlighted a significant cluster in the orbitofrontal cortex (*p_FWE_ <* 0.05, HC *>* HP), and (B) the post-hoc comparison of HPs with Non-Hospitalized Patients (NHPs) also highlighted a cluster in the orbitofrontal cortex (*p_FWE_ <* 0.05, NHP *>* HP).

### 3.4. Fractional anisotropy differences between controls and COVID-19 survivors

Tract-based spatial statistics (TBSS) analyses revealed widespread alterations in fractional anisotropy (FA) across the white matter skeleton. Compared with HCs, CRPs exhibited significant reductions in FA bilaterally within the inferior fronto-occipital fasciculus (IFOF), with a stronger effect observed on the left hemisphere, as shown in Figure 2A. Conversely, CRPs showed increased FA in the bilateral internal capsules, splenium of the corpus callosum, and left superior longitudinal fasciculus (SLF) relative to controls (Figure 2B).

**Figure 2:**
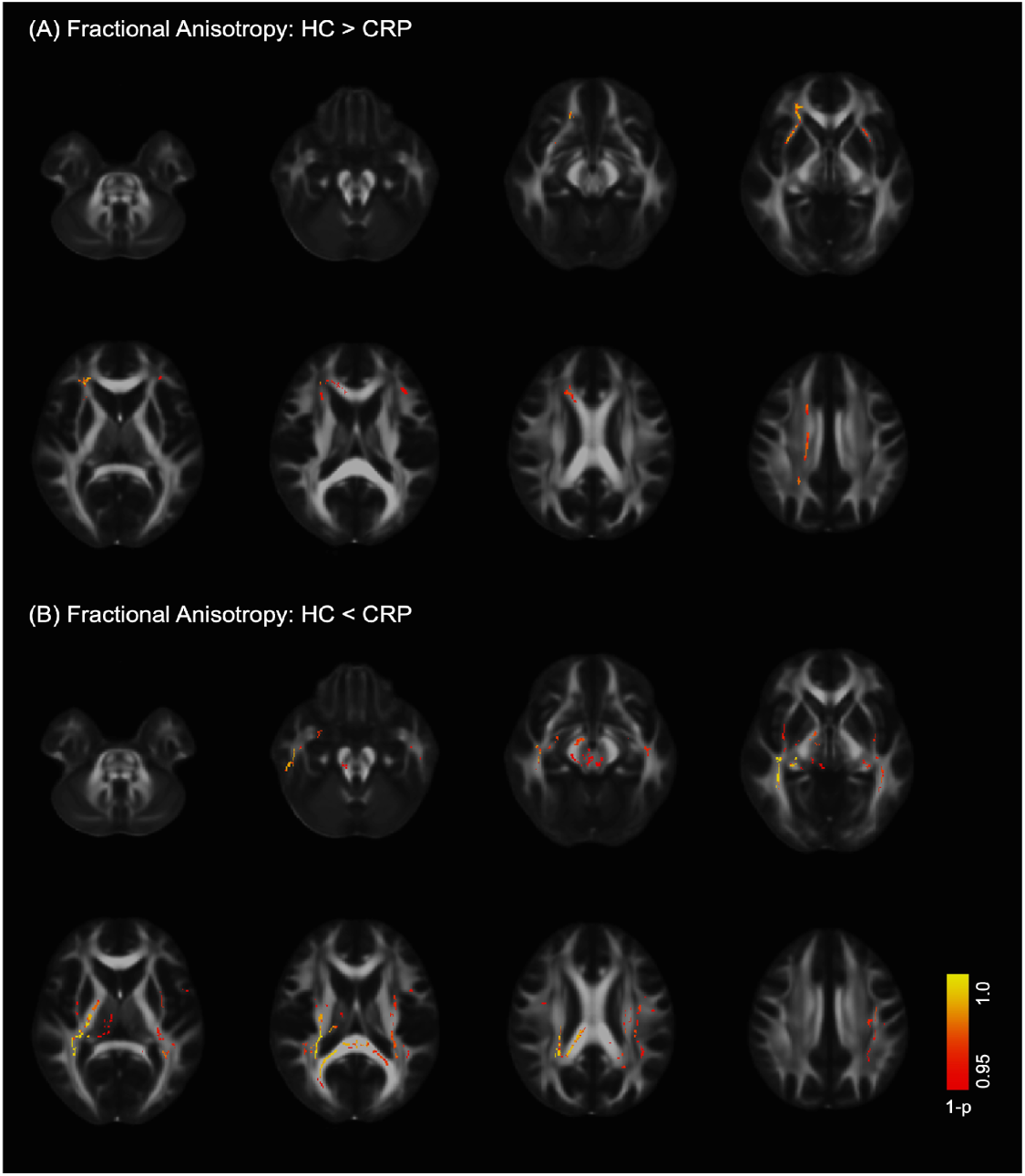
Clusters showing significant differences in Fractional Anisotropy (FA) between the Healthy Controls (HCs) and COVID-19 Recovered Patients (CRPs) using Tract-Based Spatial Statistics (TBSS) (*p_corr_ <* 0.05). (A) Significant differences in FA were observed in the right and left inferior fronto-occipital fasciculus (HC *>* CRP). (B) Significantly increased FA in CRPs was observed in the bilateral internal capsules (HC *<* CRP), the splenium of the corpus callosum (HC *<* CRP), and the left superior longitudinal fasciculus (SLF) (HC *<* CRP).

### 3.5. COVID-19 survivors show reduced mean diffusivity

We observed reduced MD in several regions of white matter in CRPs (*p_corr_ <* 0.05), including the corpus callosum splenium, the right IFOF, the external capsule of the left, the left cingulum, and the forceps major (Figure 3). Lower MD values overlapped with both areas of lower and higher FA values for the CRP group. We did not observe any regions with significantly increased MD in CRPs compared with HCs (*p_corr_ >* 0.05).

**Figure 3:**
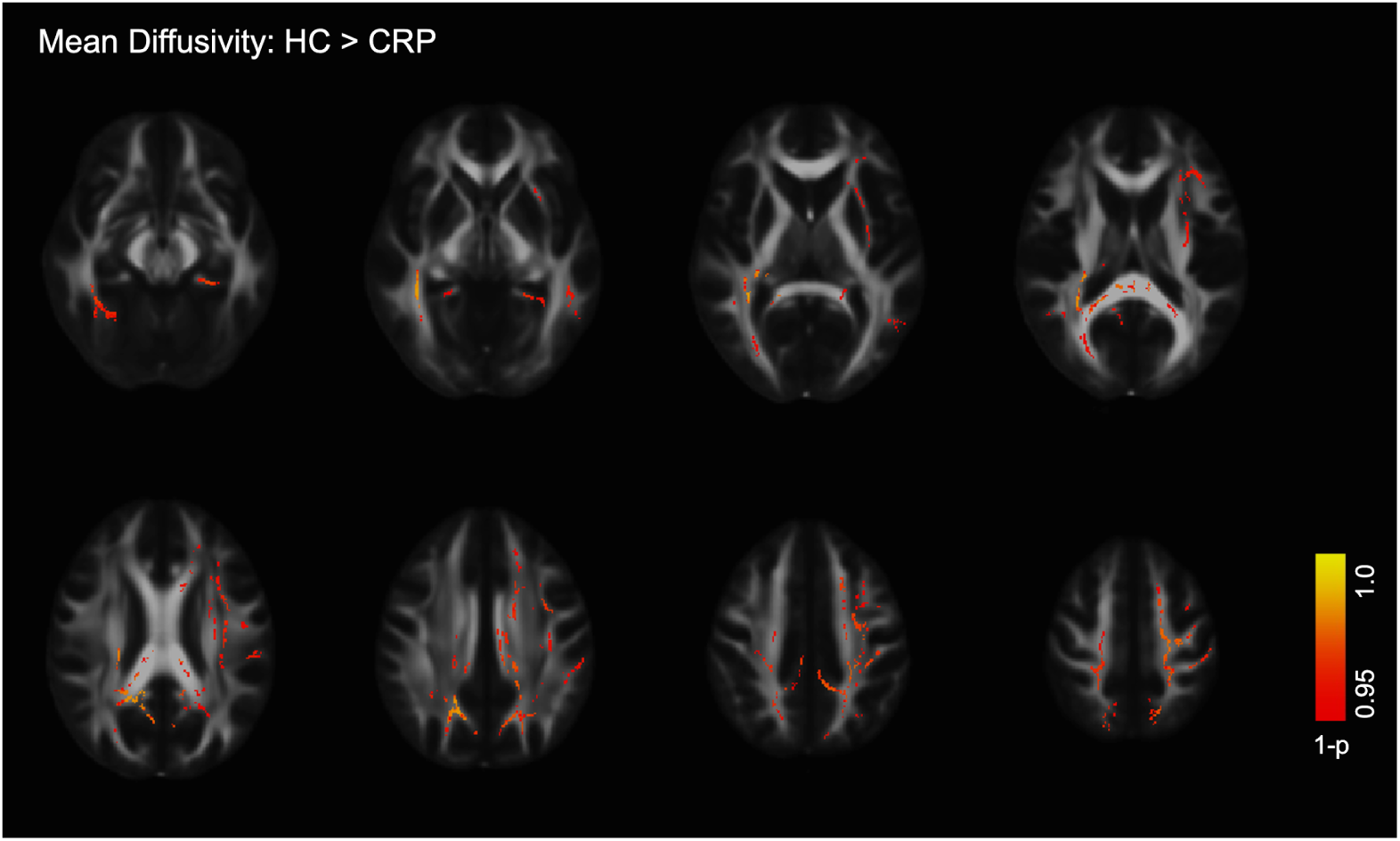
Clusters showing significant differences in Mean Diffusivity (MD) between the Healthy Controls (HCs) and COVID-19 Recovered Patients (CRPs) using Tract-Based Spatial Statistics (TBSS) (*p_corr_ <* 0.05). Significant differences in MD were observed in the splenium of the corpus callosum, right inferior fronto-occipital fasciculus, left external capsule, left cingulum, and the forceps major tract (HC>CRP).

### 3.6. Differences in mode of anisotropy in COVID-10 survivors

Comparison of MO maps between HCs and CRPs revealed that MO was significantly lower in CRPs as compared to HCs in widespread regions of white matter (*p_corr_ <* 0.05). Significant clusters were found in the uncinate fasciculus, corpus callosum, internal capsule, and cortical white matter projecting to the prefrontal and parietal cortices (Figure 4A). On the other hand, posterior parts of the corpus callosum and corona radiata showed higher MO in CRPs as compared to HCs (*p_corr_ <* 0.05) (Figure 4B).

**Figure 4:**
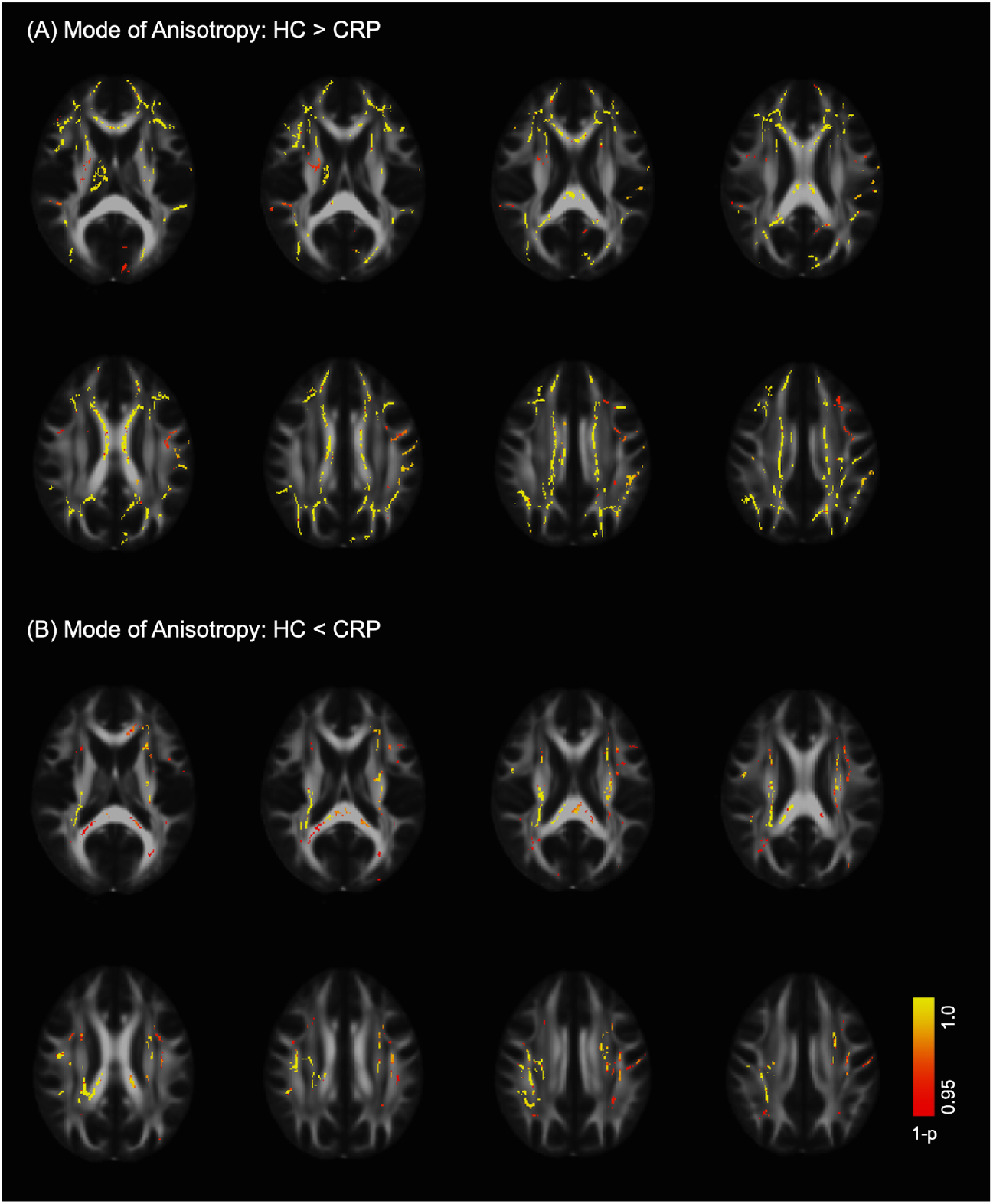
Clusters showing significant differences in mode of anisotropy (MO) between the Healthy Controls (HCs) and COVID-19 Recovered Patients (CRPs) using Tract-Based Spatial Statistics (TBSS) (*p_corr_ <* 0.05). (A) Significant differences in MO were observed in the uncinate fasciculus, corpus callosum, internal capsule, and cortical white matter projecting to the prefrontal and parietal cortices (HC>CRP).(B) Significant differences in MO (CRP>HC) were found in the posterior parts of the corpus callosum and corona radiata.

### 3.7. Fractional anisotropy abnormalities observed with severity stratification

Next, a GLM analysis was used to assess the effect of infection severity based stratification on the FA values in HCs, NHPs, and HPs. We observed an overall significant effect of stratification on the FA values among these groups (*p_corr_ <* 0.05) (see supplementary Figure S3.A). Further pairwise post-hoc tests were conducted to discern the groups that were significantly impacted.

We found clusters that showed significantly lower FA values in HPs as compared with HCs (*p_corr_ <* 0.05). The clusters are presented in Figure 5A. Key regions included the left anterior thalamic radiation and parts of the left forceps minor. Both NHPs and HPs also each exhibited clusters with higher FA than HCs (*p_corr_ <* 0.05). HPs showed regions with significantly elevated FA compared with HCs (*p_corr_ <* 0.05) in the splenium of the corpus callosum, bilateral internal capsules, superior corona radiata, left SLF, and posterior parts of IFOF (Figure 5B). NHPs showed significantly elevated FA in the forceps major, splenium of the corpus callosum, right and left IFOF, the left SLF, cingulum, and the left anterior thalamic radiation (*p_corr_ <* 0.05) compared with HCs (Figure 5C). No regions showed significantly different FA between HPs and NHPs (*p_corr_ >* 0.05).

**Figure 5:**
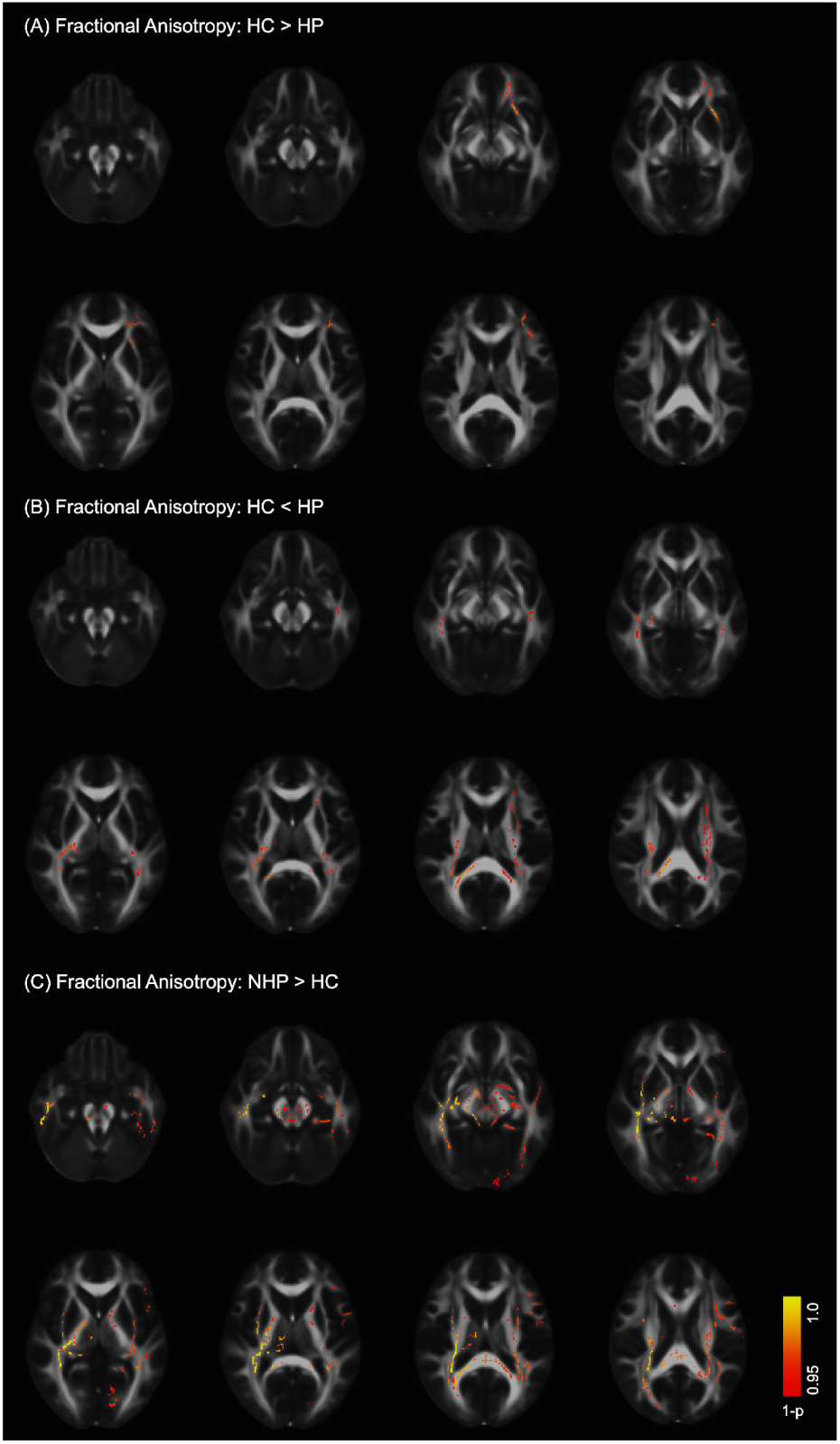
Significant clusters that highlight the effect of severity-based stratification on Fractional Anisotropy (FA) (*p_corr_ <* 0.05). Post-hoc comparisons between Healthy Controls (HCs) and Hospitalized Patients (HPs) showed (A) a significant reduction in FA in HPs in the left anterior thalamic radiation, and parts of the left forceps minor (HC>HP), and (B) an increase in FA values in HPs in the splenium of corpus callosum, internal capsules (right and left), superior corona radiata, left superior longitudinal fasciculus, and posterior parts of inferior fronto-occipital fasciculus (HC *<* HP). (C) Post-hoc comparisons between Healthy Controls (HCs) and Non-Hospitalized Patients (NHPs) showed a significant increase in FA values in NHPs in the forceps major, splenium of corpus callosum, right and left inferior fronto-occipital fasciculus, the left superior longitudinal fasciculus, cingulum, and the left anterior thalamic radiation (NHP *>* HC).

### 3.8. Mean diffusivity differences with severity stratification

The effect of infection severity-based stratification on MD values was marginally significant (*p_corr_ <* 0.12) (see supplementary Figure S3.B). However, we found significant effects when post-hoc tests were conducted. Upon pairwise comparison of MD maps, both NHPs and HPs exhibited regions where the MD values were significantly lower than for HCs (*p_corr_ <* 0.05). HPs showed significantly reduced MD values in the forceps major, left inferior longitudinal fasciculus (ILF), the left IFOF, and the left SLF (Figure 6A), while NHPs showed significantly reduced MD values in the right IFOF (Figure 6B). We did not observe a significant difference in MD values between HPs and NHPs (*p_corr_ >* 0.05).

**Figure 6:**
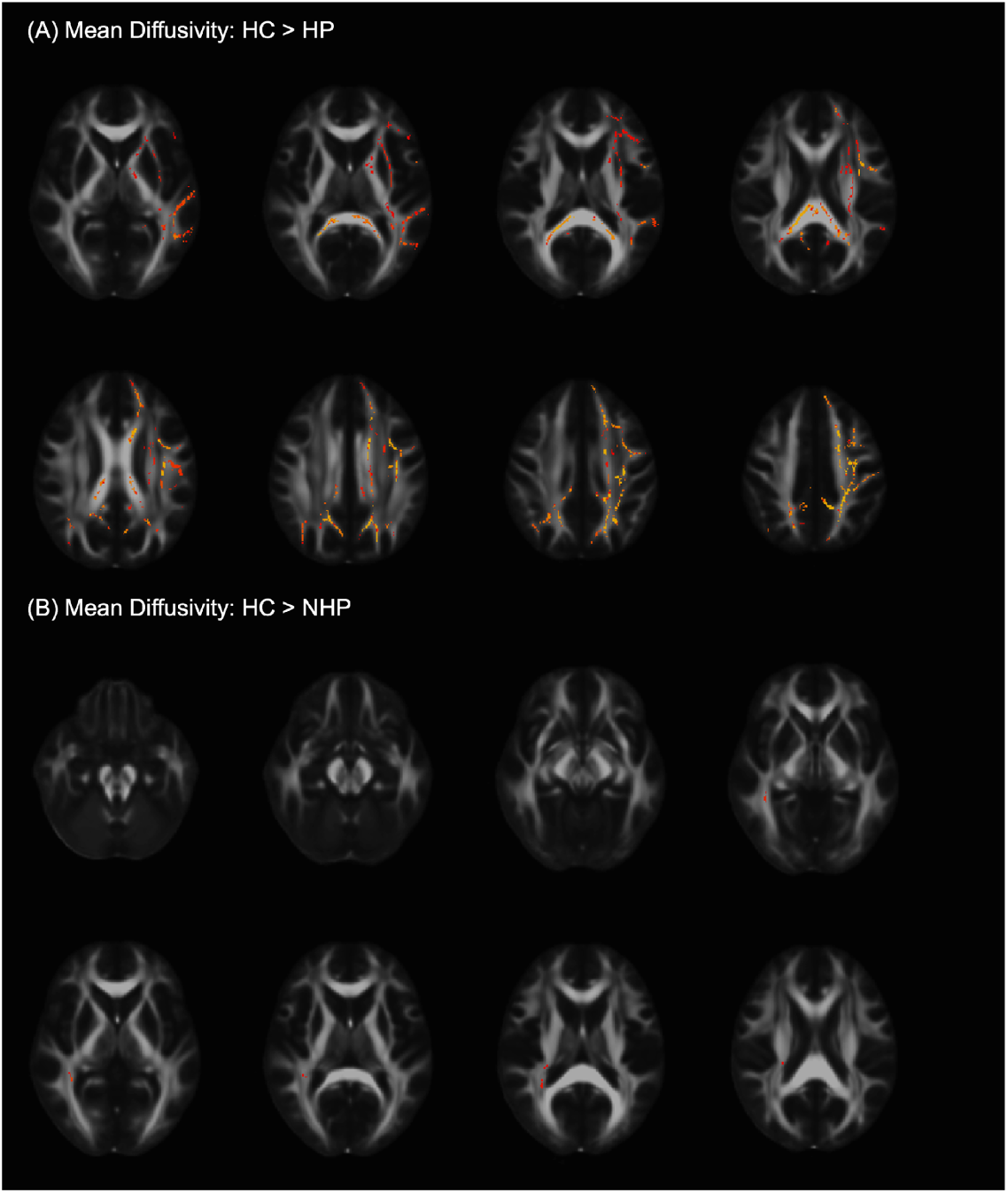
Significant clusters that highlight the effect of severity-based stratification on Mean Diffusivity (*p_corr_ <* 0.05). (A) Post-hoc comparisons between Healthy Controls (HCs) and Hospitalized Patients (HPs) showed a significant decrease in MD values in HPs in the forceps major, left inferior longitudinal fasciculus, the left inferior fronto-occipital fasciculus, and the left superior longitudinal fasciculus (HC *>* HP). (B) Post hoc comparisons between Healthy Controls (HCs) and Non-Hospitalized Patients (NHPs) showed a significant decrease in MD values in NHPs in the right inferior fronto-occipital fasciculus (HC *>* NHP).

### 3.9. Effect of severity stratification on mode of anisotropy

We found a significant effect of severity-based stratification on MO (*p_corr_ <* 0.05) (see supplementary Figure S3.C). Post hoc comparison revealed that MO was significantly lower for HPs as compared to HCs in the left posterior thalamic radiation, external capsule, and the crossings of uncinate fasciculus and anterior thalamic radiations (*p_corr_ <* 0.05) (Figure S4.A). NHPs showed lower values of MO than HCs in the corpus callosum, anterior thalamic radiations, corticospinal tract, and white matter projections in the posterior parietal cortex (*p_corr_ <* 0.05) (Figure S4.B). HPs showed higher MO values than HCs in the corticospinal tract and parts of the splenium of corpus callosum (*p_corr_ <* 0.05) (Figure S4.C). NHPs showed higher MO values than HCs in the inferior fronto-occipital fasciculus and the corticospinal tract (*p_corr_ <* 0.05) (Figure S4.D). No significant differences were observed upon post hoc comparison of NHPs and HPs (*p_corr_ >* 0.05).

### 3.10. No significant differences in thalamocortical FC in COVID-19 survivors and controls

We compared the TC-FC maps for each thalamic seed across the two cohorts, HCs and CRPs. The results were obtained using a two-sample t-test, and significant clusters were identified by using a height threshold of *p_unc_ <* 0.001, and corrected for multiple comparisons at a family-wise error (FWE) of *p_FWE_ <* 0.05. We did not observe any significant differences between the TC-FC of HCs and CRPs across the three thalamic nuclei tested (*p_FWE_ <* 0.05).

### 3.11. Effect of severity-based stratification on functional connectivity with mediodorsal thalamic nuclei

To test the effect of infection severity-based stratification on the FC of the brain with various thalamic nuclei, we used the HC, NHP, and HP cohorts. We observed a significant effect of stratification on TC-FC in the case of the left and right MDN (*p_FWE_ <* 0.05) (see supplementary Figure S5). No other thalamic seeds showed a significant effect of stratification (*p_FWE_ <* 0.05). Further post-hoc tests were conducted to identify the regions that were impacted.

With the seed in the left MDN, we found significant clusters in multiple pairwise post-hoc comparisons (see Figure 7). First, the group comparison between HCs and HPs highlighted three clusters with significant differences (*p_FWE_ <* 0.05, HC < HP) (Figure 7A), in the (i) left inferior visual association cortex, (ii) right supramarginal gyrus, angular gyrus, and fusiform gyrus, and (iii) right visual association cortex. Comparison of the NHPs and HP yielded three clusters with significant differences (*p_FWE_ <* 0.05) (Figure 7B): the first cluster was observed in the right superior parietal cortex, near the secondary somatosensory cortex, the second in the right angular gyrus close to the supramarginal gyrus, and the third cluster was found in the left cuneal cortex. Finally, the comparison of HCs and NHPs revealed a cluster in the left anterior cingulate cortex (*p_FWE_ <* 0.05, NHP > HC) (Figure 7C).

**Figure 7:**
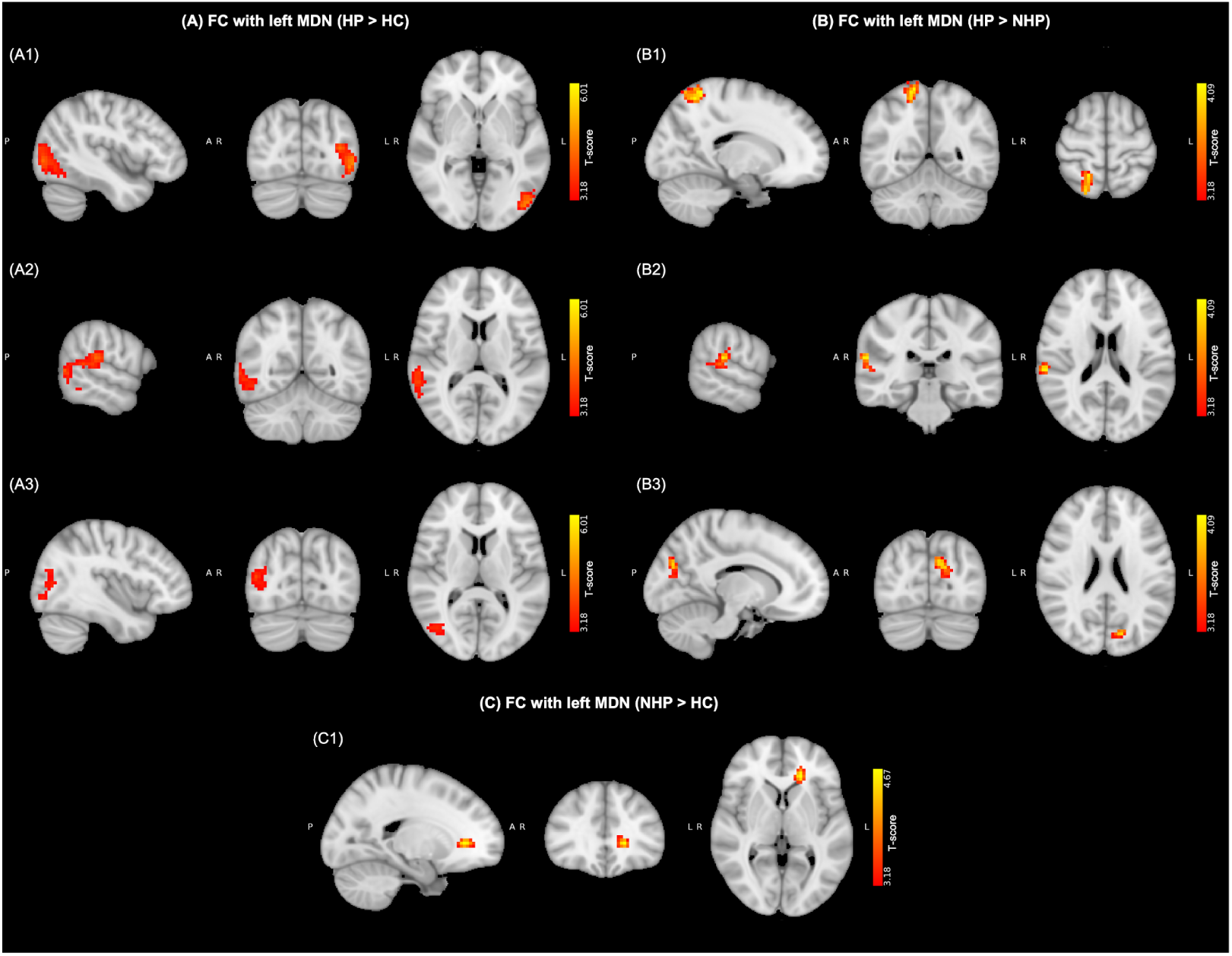
Regions that exhibit significant differences in functional connectivity (FC) with the left Medio-dorsal nucleus (MDN) upon comparing Healthy Controls (HCs), Non-Hospitalized Patients (NHPs), and Hospitalized Patients (HPs) (*p_FWE_ <* 0.05). (A) Clusters with increased FC in HPs (HP *>* HC) were observed in the (A1) left inferior visual association cortex, (A2) right supramarginal gyrus, angular gyrus, and fusiform gyrus, and (A3) right visual association cortex. (B) Clusters with increased FC in HPs (HP *>* NHP) were observed in the (B1) right superior parietal cortex, (B2) right angular gyrus, and (B3) left cuneal cortex. (C) A cluster with increased FC in NHPs (NHP *>* HC) was observed in the left anterior cingulate cortex.

Furthermore, when the seed was placed in the right MDN, we observed significant clusters in multiple post-hoc tests (see Figure 8). The comparison of HPs with HCs yielded three significant clusters (*p_FWE_ <* 0.05, HC < HP, Figure 8A), localized in the (i) left inferior visual association cortex near the fusiform gyrus, (ii) right fusiform gyrus extending to the angular gyrus, and (iii) bilateral primary visual cortex. Comparison of NHPs and HPs highlighted significant clusters (*p_FWE_ <* 0.05, NHP < HP, Figure 8B) in the (i) right superior parietal cortex near the secondary somatosensory cortex, and (ii) medial left dorsal visual association cortex. Comparison between NHPs with HCs yielded similar results to the contralateral MDN, with a significant cluster in the left anterior cingulate cortex (*p_FWE_ <* 0.05, HC < NHP, Figure 8C). A summary of the clusters obtained is provided in Table 2.

**Figure 8:**
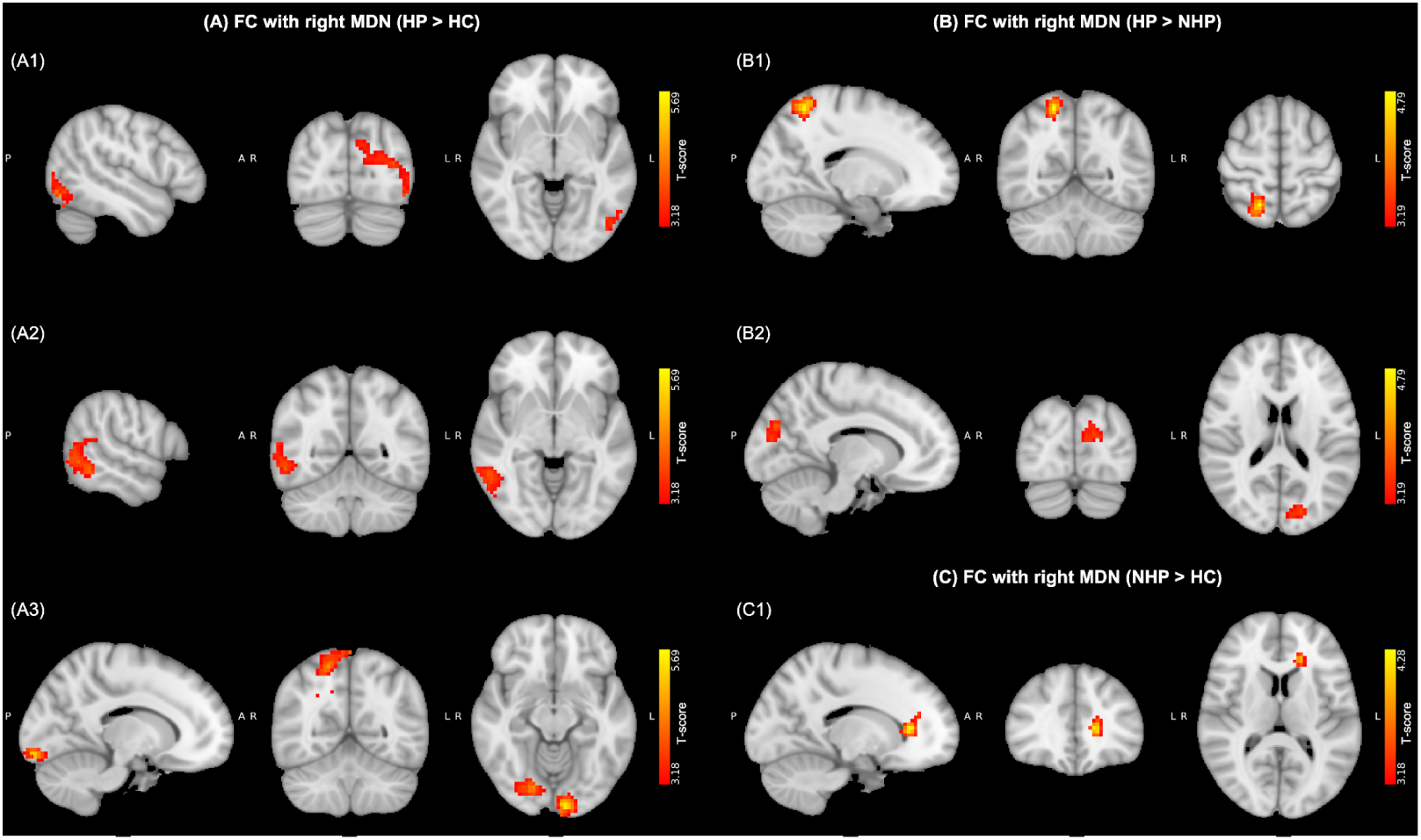
Regions that exhibit significant differences in functional connectivity (FC) with the right Medio-dorsal nucleus (MDN) upon comparing Healthy Controls (HCs), Non-Hospitalized Patients (NHPs), and Hospitalized Patients (HPs) (*p_FWE_ <* 0.05). (A) Clusters with increased FC in HPs (HP *>* HC) were observed in the (A1) left inferior visual association cortex, (A2) right fusiform gyrus, and (A3) bilateral primary visual cortex. (B) Clusters with increased FC in HPs (HP *>* NHP) were observed in the (B1) right superior parietal cortex, and (B2) medial left dorsal visual association cortex. (C) A cluster with increased FC in NHPs (NHP *>* HC) was observed in the left anterior cingulate cortex.

**Table 2:**
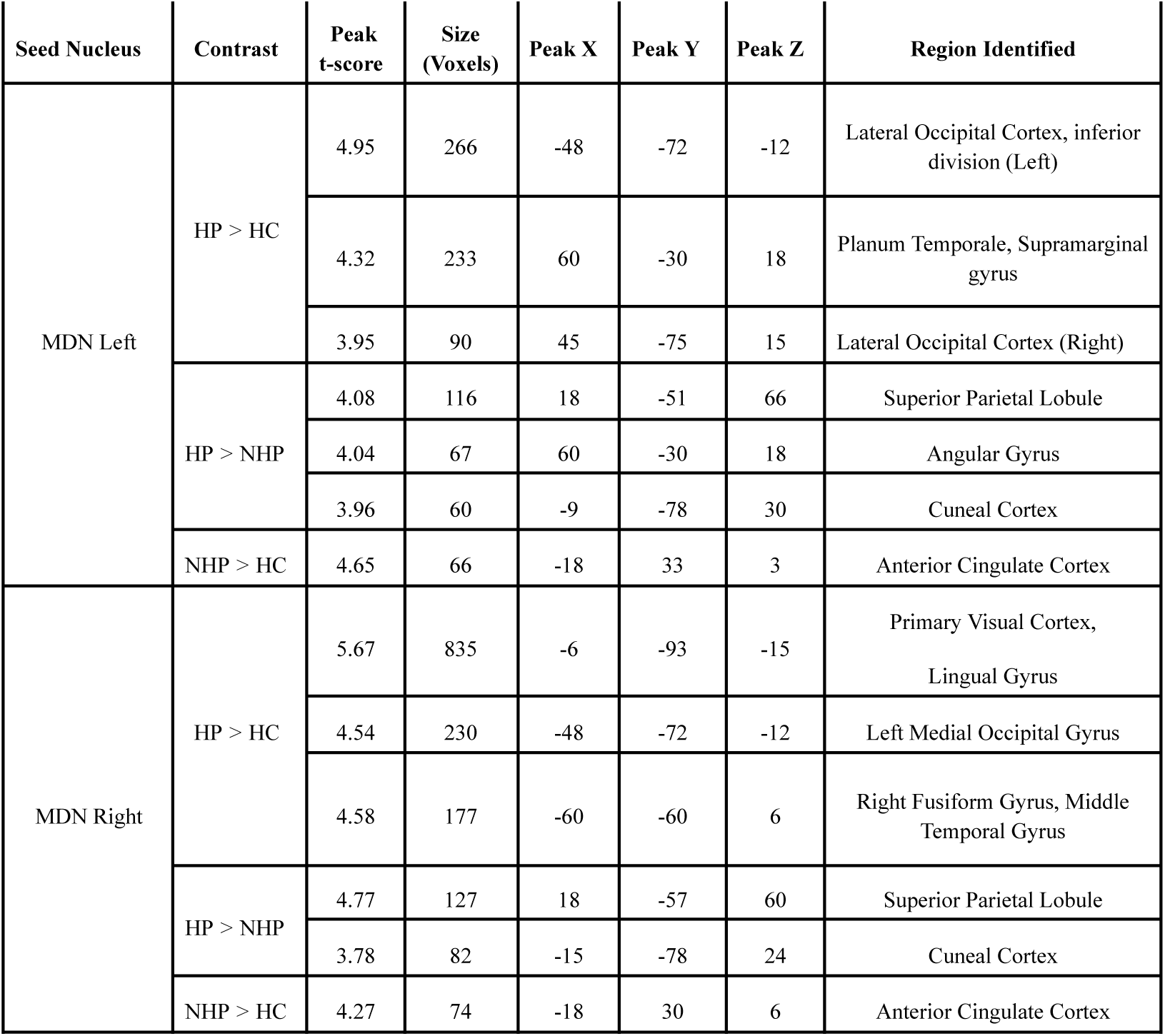
Summary of clusters observed showing significant differences in thalamocortical functional connectivity among the Healthy Controls (HCs), and the COVID-19 Recovered Non-Hospitalized Patients (NHPs), and Hospitalized Patients (HPs). Key: MDN = Medio-Dorsal Nucleus.

| Seed Nucleus | Contrast | Peak t-score | Size (Voxels) | Peak X | Peak Y | Peak Z | Region Identified |
| --- | --- | --- | --- | --- | --- | --- | --- |
| MDN Left | HP > HC | 4.95 | 266 | -48 | -72 | -12 | Lateral Occipital Cortex, inferior division (Left) |
|  |  | 4.32 | 233 | 60 | -30 | 18 | Planum Temporale, Supramarginal gyrus |
|  |  | 3.95 | 90 | 45 | -75 | 15 | Lateral Occipital Cortex (Right) |
|  | HP > NHP | 4.08 | 116 | 18 | -51 | 66 | Superior Parietal Lobule |
|  |  | 4.04 | 67 | 60 | -30 | 18 | Angular Gyrus |
|  |  | 3.96 | 60 | -9 | -78 | 30 | Cuneal Cortex |
|  | NHP > HC | 4.65 | 66 | -18 | 33 | 3 | Anterior Cingulate Cortex |
| MDN Right | HP > HC | 5.67 | 835 | -6 | -93 | -15 | Primary Visual Cortex, Lingual Gyrus |
|  |  | 4.54 | 230 | -48 | -72 | -12 | Left Medial Occipital Gyrus |
|  |  | 4.58 | 177 | -60 | -60 | 6 | Right Fusiform Gyrus, Middle Temporal Gyrus |
|  | HP > NHP | 4.77 | 127 | 18 | -57 | 60 | Superior Parietal Lobule |
|  |  | 3.78 | 82 | -15 | -78 | 24 | Cuneal Cortex |
|  | NHP > HC | 4.27 | 74 | -18 | 30 | 6 | Anterior Cingulate Cortex |

## 4. Discussion

This study used multimodal MRI to examine structural, microstructural, and functional brain alterations in individuals recovered from COVID-19. We investigated whether (1) long-term changes persist in grey or white matter, (2) these effects vary with disease severity, and whether (3) thalamocortical connectivity shows evidence of reorganisation. Three main observations emerged. First, severity-related grey-matter loss was confined to the orbitofrontal cortex and frontal pole in hospitalized patients. Second, diffusion imaging revealed widespread, partly severity-related microstructural alterations with both decreases and paradoxical increases in fractional anisotropy (FA), accompanied by reduced mean diffusivity (MD) and complex changes in mode of anisotropy (MO). Third, resting-state functional MRI showed altered thalamocortical connectivity exclusively in the mediodorsal nucleus (MDN), with increased coupling to anterior cingulate, parietal, and occipital cortices. Together, these findings point to enduring perturbation of fronto-limbic and thalamocortical networks after COVID-19, with the strongest effects in those who experienced severe illness.

### 4.1. Orbitofrontal grey-matter loss and severity effects

Our VBM analysis revealed no whole-group differences between CRPs and HCs but identified focal orbitofrontal thinning in HPs. This pattern mirrors longitudinal findings from the UK Biobank, which showed cortical thinning and tissue-contrast loss in orbitofrontal and olfactory-limbic regions following infection [5]. Importantly, the same study noted that thalamic differences were largely driven by pre-infection variation, suggesting that individuals with lower baseline thalamic integrity may have been more susceptible to infection or severe systemic response. In this context, our observation of preserved thalamic volume but altered thalamo-cortical coupling may reflect compensatory adaptation within an already vulnerable network.

Involvement of the orbitofrontal cortex (OFC) aligns with both behavioural and imaging evidence linking this region to anosmia, emotional regulation, and motivation; all frequently disrupted in post-COVID syndrome [44]. OFC thinning may underlie combined effects of olfactory deafferentation and neuroinflammatory or vascular injury. In our earlier work, we identified consistent basal-ganglia and limbic abnormalities [16] and tract alterations within the fornix and cingulum [14], situating the present OFC finding within a broader fronto-limbic pattern of disruptions.

### 4.2. White-matter microstructure: widespread but heterogeneous change

TBSS revealed extensive alterations in white-matter integrity across association and commissural tracts, including the IFOF, SLF, cingulum, and callosal fibres. Reduced MD was the dominant feature, consistent with restricted water diffusion due to inflammatory cellular infiltration or cytotoxic oedema [45, 46]. Similar DTI signatures have been described in post-infectious neuroinflammatory states and in longitudinal COVID-19 cohorts months after recovery [6, 8].

A notable aspect of our findings is the paradoxical increase in FA in certain regions, particularly within the internal capsule and splenium, which co-occurred with increases in MO. The combination of high FA and high MO implies loss of crossing-fibre populations, leading to greater directional coherence along surviving tracts rather than genuine microstructural improvement [28]. This interpretation reconciles the mixed changes in FA seen across studies, i.e., both decreased and increased FA can reflect pathological change depending on the geometry of affected fibres. The coexistence of reduced MD and elevated FA/MO in posterior callosal regions therefore supports a process of partial fibre loss or selective degeneration within complex fibre bundles rather than homogeneous axonal injury.

Severity-dependent differences, more prominently in HPs, further support a dose-response relationship between acute disease burden and subsequent white-matter remodelling, aligning with reports that lower oxygenation and higher inflammatory load predict more extensive diffusion abnormalities [10]. These microstructural findings converge with our earlier fixel-based analysis in the same cohort [15], strengthening the evidence that limbic-cortical tracts are persistently altered after infection.

### 4.3. Nucleus-specific thalamocortical dysfunction

At the functional level, we observed no global differences in TC-FC between groups, but severity-stratified analysis revealed increased connectivity from the MDN to the anterior cingulate, supramarginal gyrus, and visual cortex. Notably, the pulvinar and ventrolateral nuclei showed no significant alterations. The MDN’s selective involvement is physiologically coherent. It is reciprocally connected with the olfactory and orbitofrontal cortices [47, 48], regions prominently affected in COVID-19 due to viral entry via the olfactory epithelium [49, 50]. This anatomical proximity offers a plausible pathway for trans-synaptic or inflammatory propagation influencing MDN circuits.

Our results are aligned with Leitner et al. [22], who reported increased MDN connectivity with prefrontal and parietal cortices in post-COVID patients experiencing fatigue, and complement observations of thalamic involvement in cognitive dysfunction across other post-viral syndromes [51]. Increased MDN connectivity may therefore represent compensatory up-regulation of thalamic relay function in response to cortical inefficiency or subtle OFC injury. Alternatively, it could reflect maladaptive hyperconnectivity driven by persistent glial activation or altered inhibitory gating within thalamo-cortical loops [18]. The presence of this effect in both NHPs and HPs (stronger in the HPs) supports a continuum of network-level adaptation proportional to acute disease severity.

Importantly, the absence of effects in pulvinar and ventrolateral nuclei highlights the functional specificity of thalamic involvement. The pulvinar nucleus primarily mediates visuospatial attention and the ventrolateral nucleus is involved in motor integration, domains less consistently affected in PCS [52]. The selective disruption in TC-FC of the MDN therefore reinforces the idea of fronto-limbic–thalamic dysregulation as a central axis of post-COVID brain change [53].

### 4.4. Integrating modalities

Across modalities, our data reveal converging disruption within a fronto-limbic–thalamic circuit encompassing the OFC, limbic white-matter tracts, and mediodorsal thalamus. OFC volume loss may signal cortical degeneration or deafferentation, white-matter alterations in the IFOF and cingulum indicate impaired cortico-subcortical communication, and increased MDN connectivity may represent either a compensatory response or a sign of ongoing thalamic dysregulation. The overall picture is one of network-level disruption rather than isolated lesion effects, consistent with PCS as a disorder of distributed brain systems.

While some studies report reduced thalamic or cortical connectivity post COVID [13], others, including Leitner et al. [22], describe increased coupling. Our results help reconcile this divergence by showing that group averaging can mask severity-dependent effects. Our results show that once stratified, both directions of change can coexist across nuclei and participants. Similarly, diffusion studies alternately report increased or decreased FA in PCS. Accounting for MO variations and tract geometry clarifies these apparent inconsistencies. Together, these findings emphasise the need for stratified, multimodal designs when assessing post-COVID neural alterations.

## 5. Limitations

Our study was cross-sectional in nature and therefore it is difficult to comment on causality. The hospitalised subgroup in our cohort was relatively small, and infection-to-scan intervals varied. MRI acquisitions at 3T with single shell diffusion limited microstructural modelling. Future work should employ multi-shell and longitudinal approaches to study these aspects. Although we controlled for head motion and global signal in fMRI, residual physiological confounds may influence functional connectivity estimates. Finally, hospitalisation was used as a proxy for severity which might be confounded by pre-existing susceptibilities and health concerns. Instead incorporating oxygen saturation, inflammatory markers, and cognitive outcomes would refine interpretation of variations with respect to infection severity.

## 6. Implications and Future Directions

Despite these limitations, this study provides cohesive multimodal evidence that COVID-19 leads to enduring perturbations of fronto-limbic and thalamo-cortical networks that vary with infection severity. The selective dysfunction in the MDN highlights the role of the thalamus as a vulnerable and integrative hub, potentially both a predisposing factor (as suggested by pre-infection differences in UK Biobank data) and a target of secondary reorganisation. These findings highlight vital imaging markers, i.e., OFC volume, integrity in limbic tracts, and MDN connectivity, that could inform longitudinal tracking or targeted interventions.

Future research should integrate longitudinal imaging with symptom and cognitive assessment to test whether these markers predict recovery or persistence of fatigue and cognitive dysfunction. Advanced diffusion models (e.g. multi-shell models, free-water imaging) and vascular measures (ASL, cerebrovascular reactivity) may disentangle neuroinflammatory from neurovascular components. Finally, understanding whether targeting TC-FC through behavioural or neuromodulatory interventions improves clinical outcomes could open translational avenues for managing post-COVID cognitive fatigue.

## Data Availability

The dataset will be made available upon reasonable request to the corresponding authors, subject to institutional data-sharing policies.

