## Supplementary files for "Fronto-limbic and Thalamocortical Network Alterations after COVID-19 Recovery: a Multimodal MRI Study"

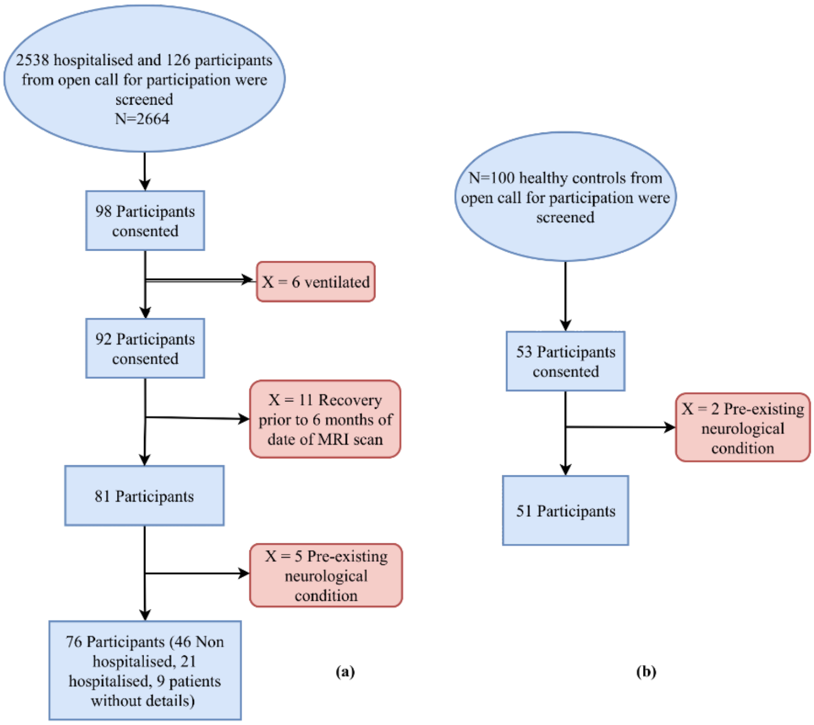


Figure S1: Flowchart of the recruitment of participants in the (a) COVID-19 Recovered Patients (CRPs) and (b) Healthy Controls (HCs) groups, following all the inclusion and exclusion criteria.

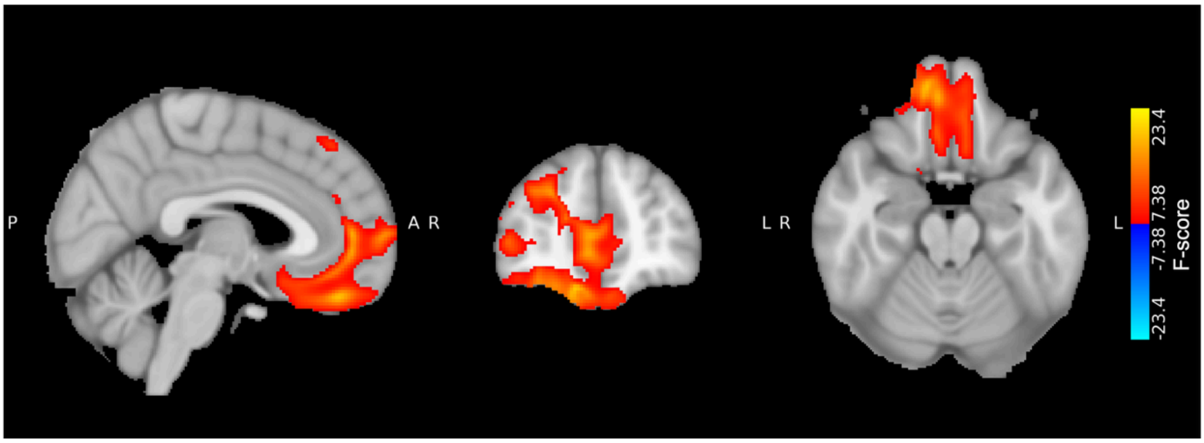


Figure S2. Clusters showing significant effect of severity-based stratification (HC vs. NHP vs. HP) on grey-matter volume derived using voxel-based morphometry. ($p_{FWE}<0.05$)

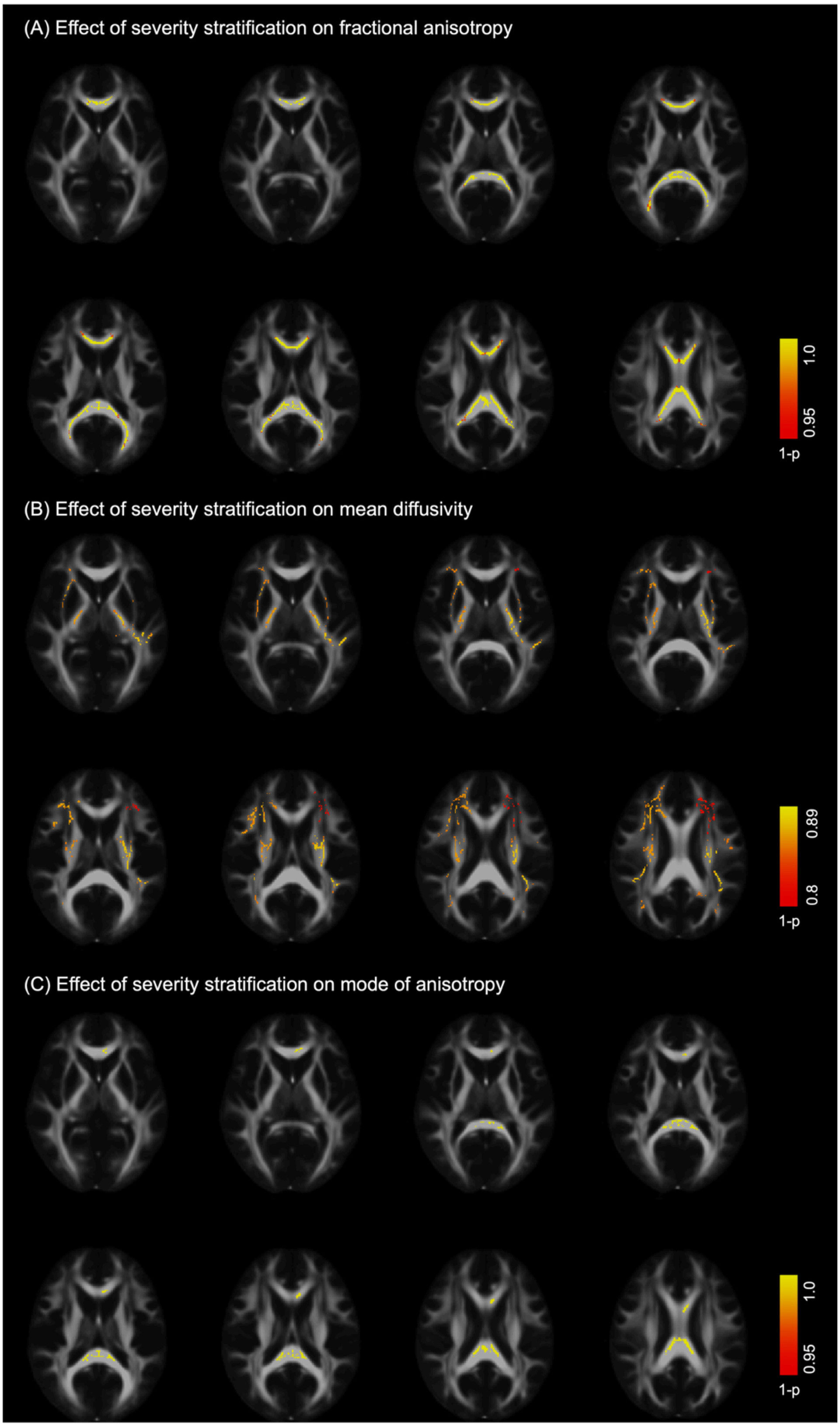


Figure S3. Clusters showing significant effect of severity-based stratification (HC vs. NHP vs. HP) on white-matter microstructure derived using tract-based spatial statistics. Effect of stratification on (A) fractional anisotropy values ($p_{corr}$<0.05), (B) mean diffusivity values($p_{corr}$<0.12), and (C) mode of anisotropy ($p_{corr}$<0.05)

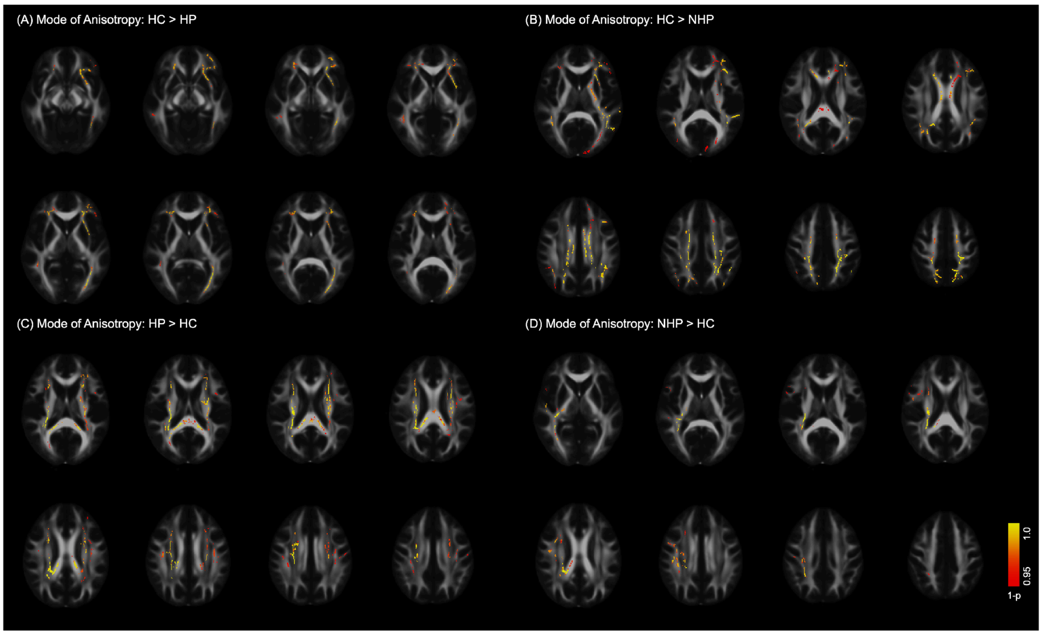


Figure S4: Significant clusters that highlight the effect of severity-based stratification on mode of anisotropy ($p_{corr} < 0.05$). (A) Post hoc comparison revealed that MO was significantly lower for HPs as compared to HCs in the left posterior thalamic radiation, external capsule, and the crossings of uncinate fasciculus and anterior thalamic radiations (HC>HP). (B) NHPs showed lower values of MO than HCs in the corpus callosum, anterior thalamic radiations, corticospinal tract, and white matter projections in the posterior parietal cortex (HC>NHP). (C) HPs showed higher MO values than HCs in the corticospinal tract and parts of the splenium of corpus callosum (HP>HC). (D) NHPs showed higher MO values than HCs in the inferior fronto-occipital fasciculus and the corticospinal tract (NHP>HC).

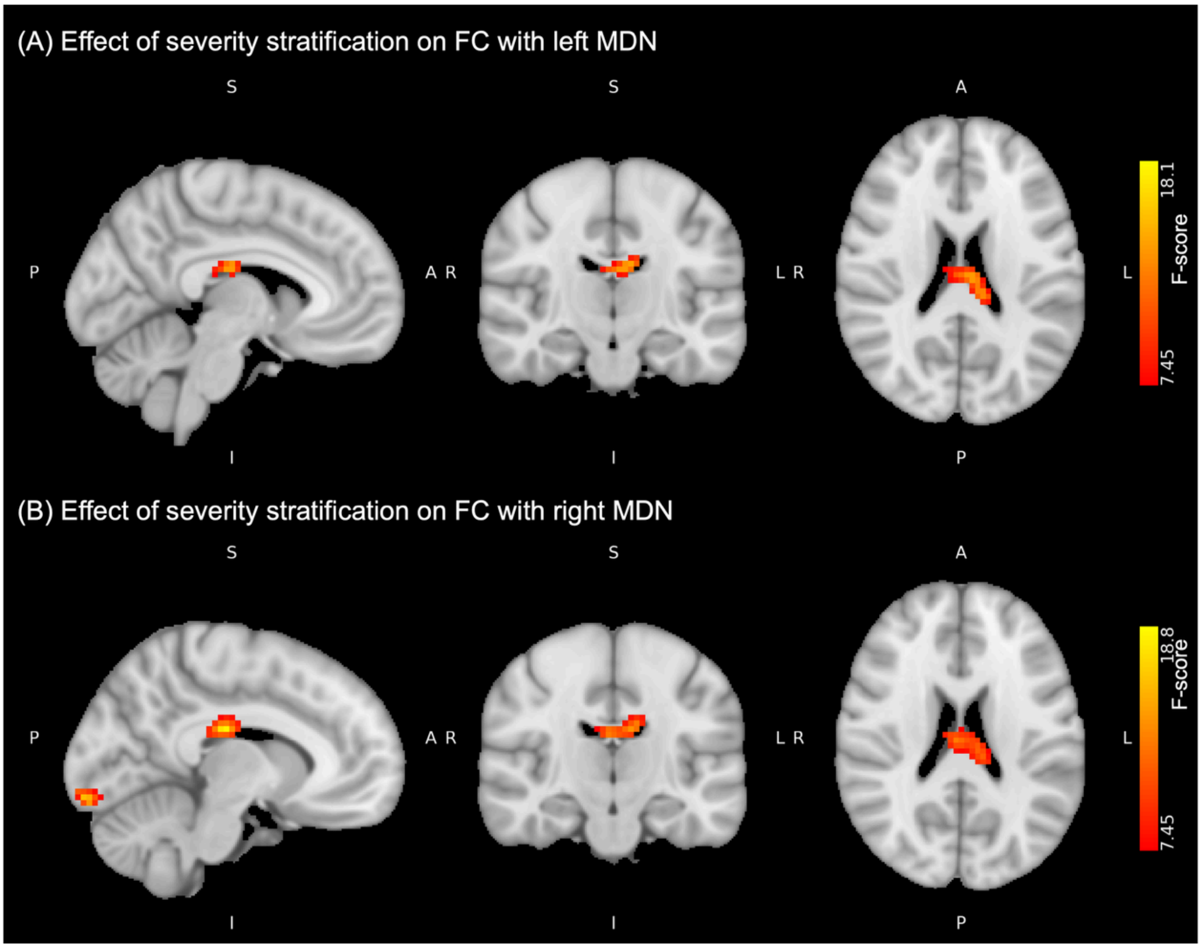


Figure S5. Clusters showing significant effect of severity-based stratification (HC vs. NHP vs. HP) on thalamo-cortical functional connectivity with the (A) left mediodorsal nucleus (MDN) ($p_{corr}$<0.05), and (B) right MDN ($p_{corr}$<0.05)
